# Resilience to dominant genetic disease in the healthy elderly

**DOI:** 10.1101/19006932

**Authors:** Paul Lacaze, Robert Sebra, Moeen Riaz, Jane Tiller, Jerico Revote, James Phung, Emily J Parker, Suzanne G Orchard, Jessica E Lockery, Rory Wolfe, Maya Strahl, Ying C Wang, Rong Chen, Daniel Sisco, Todd Arnold, Bryony A Thompson, Daniel D Buchanan, Finlay A Macrae, Paul A James, Walter P Abhayaratna, Trevor J Lockett, Peter Gibbs, Andrew M Tonkin, Mark R Nelson, Christopher M Reid, Robyn L Woods, Anne M Murray, Ingrid Winship, John J McNeil, Eric Schadt

**Affiliations:** Department of Epidemiology and Preventive Medicine, School of Public Health and Preventive Medicine, Monash University, Melbourne, VIC, Australia; Department of Genetics and Genomic Sciences, Icahn Institute for Data Science and Genomic Technology, Icahn School of Medicine at Mount Sinai, New York, USA; Sema4, Stamford, CT, USA; Department of Genomic Medicine; Family Cancer Clinic, Department of Medicine, Department of Pathology, Royal Melbourne Hospital, University of Melbourne, Parkville, VIC, Australia; Colorectal Oncogenomics Group, Department of Clinical Pathology, University of Melbourne, VIC, Australia; College of Health and Medicine, the Australian National University, Canberra, Australia; CSIRO Health and Biosecurity, North Ryde, Australia; The Walter and Eliza Hall Institute of Medical Research, Parkville, VIC, Australia; Menzies Institute for Medical Research, University of Tasmania, Hobart, TAS, Australia; School of Public Health, Curtin University, Perth, WA, Australia; Berman Center for Outcomes and Clinical Research, Hennepin Healthcare Research Institute, Hennepin Healthcare, Minneapolis, MN, USA

## Abstract

Here we describe genomic screening of the healthy elderly to identify those resilient to adult-onset genetic disease, despite being at exceptionally high genetic risk. We sequenced 13,131 individuals aged 70 or older (mean age 75 years) from the ASPirin in Reducing Events in the Elderly (ASPREE) trial. Participants had no prior history of cardiovascular disease, life-threatening cancer, persistent physical disability or dementia. We compared the prevalence of pathogenic variants in medically actionable autosomal dominant disease genes with that from the UK Biobank population, and assessed their clinical impact using personal medical history and adjudicated study outcomes during 4.5 years of follow-up. The frequency of pathogenic variants was less than reported among the younger UK Biobank population, suggesting these variants confer a survival disadvantage during the middle years of life. Yet we identified 141 individuals with pathogenic variants free of any associated disease up to average age 79.5 years. Further study of these elderly resilient individuals might help uncover genetic mechanisms that protect against the development of disease.

## INTRODUCTION

Few genetic studies focus on the healthy elderly (1-6). By contrast, most genetic studies ascertain participants based on clinical presentation and/or family history of genetic disease (7-11). The healthy elderly, especially those who remain unaffected despite carrying genetic risk factors, are uniquely valuable. These resilient individuals may be enriched for protective genetic factors, which if understood, could inform the design of novel preventive or therapeutic strategies (12).

Previously, we identified rare exceptional ‘resilience candidates’ with protection against severe Mendelian childhood disorders by analysis of 589,306 genomes (13). Here, we take a new approach to identify individuals resilient to adult-onset genetic disease by direct screening of a healthy elderly cohort (14). As a means of identifying factors that protect against disease in resilient individuals, screening the healthy elderly provides a path to employ statistical approaches for their identification (15). The number of individuals in healthy elderly populations resilient to adult-onset disease is significantly higher than those resilient to childhood-onset disorders, where extremely large numbers need to be screened to identify small numbers of resilient individuals (13).

To estimate the rate at which individuals from a healthy elderly population are resilient to adult-onset, autosomal dominant disease, despite carrying pathogenic variants for a lifetime, we sought to screen 13,131 healthy elderly individuals enrolled in the ASPirin in Reducing Events in the Elderly (ASPREE) trial (16-18). We focused on screening for pathogenic variants in dominant genes of high clinical interest, defined as medically actionable by the American College of Medical Genetics and Genomics (ACMG) (19). Sequencing of participants aged minimum 70 years (mean age 75 years) is accompanied by documented outcome assessment from a clinical trial, to confirm the presence or absence of associated disease.

## METHODS

### STUDY POPULATION

All participants were enrolled in the ASPirin in Reducing Events in the Elderly (ASPREE) study, a randomized, placebo-controlled trial for daily low-dose aspirin (16-18). Study design (20), recruitment (21), baseline characteristics (22) and results (16-18) have been published previously. Genetic analysis was conducted on 13,131 samples from Australian participants aged 70 years or older at enrolment, free from cardiovascular disease, dementia diagnosis, physical disability, or serious illness likely to cause death within 5 years (ASPREE eligibility criteria in Supplementary Appendix).

### DNA SEQUENCING AND VARIANT ANALYSIS

A custom panel of genes with Mendelian inheritance and disease-association was designed, including 55 dominant genes defined as medically actionable by the American College of Medical Genetics and Genomics (ACMG) (19). Following standard protocols, DNA was extracted and sequenced using the Thermo Fisher Scientific S5TM XL system, to average 200X depth (see Supplementary Appendix). Variants with ‘pathogenic’ or ‘likely pathogenic’ annotation (23) and/or high-confidence predicted loss-of-function in coding regions (24) were curated following ACMG/AMP Standards and Guidelines for the Interpretation of Sequence Variants (25), including review by laboratory scientists and a clinical geneticist. Analysis was restricted to single nucleotide variants and small insertions/deletions. Variants of uncertain significance or conflicting interpretations of pathogenicity were excluded.

### PHENOTYPES AND CLINIAL OUTCOME ASSESSMENT

Available disease outcomes from the ASPREE study associated with autosomal dominant genetic conditions are summarized in Table S1. This includes self-reported medical history at enrolment (cancer history by type) and study endpoints collected during mean 4.5 years follow-up (to mean age 79.5 years). Adjudicated secondary endpoints included cancers (by type), cardiovascular events (stroke, coronary heart disease, myocardial infarction, hospitalisation for heart failure) and all-cause mortality. Coding and adjudication of endpoints has been described previously (16-18).

### DISEASE ASSOCIATION AND IDENTIFICATION OF RESILIENT CASES

We compared the prevalence of pathogenic variants in autosomal dominant genes in ASPREE versus the UK Biobank, a younger population of 49,960 adults aged average 58 years recruited from the UK general population (26). We grouped dominant genes together into related groups, then calculated the relative risk of associated disease per group in ASPREE variant carriers and 12,936 non-carriers in the trial, using personal medical history and prospective diagnoses during follow-up. We identified all pathogenic variant carriers with no evidence of associated disease symptoms to mean age 79.5 years (resilience cases).

## RESULTS

### PARTICIPANTS

The characteristics of the 13,131 sequenced participants are shown in Table 1. The mean age at enrolment was 75 years and most were White/Caucasian. Participants had no personal history of cardiovascular events at entry to the trial (16, 22). Participants with personal cancer history were not excluded from enrolment, other than those with cancer diagnoses deemed likely to cause death within five years (22).

**Table 1.** Characteristics of Sequenced Participants at Enrolment.

|  | **ASPREE**<br>**N=13,131**<br>**(mean age 75 years)** |
| --- | --- |
| **Female sex** – no. (%) | 7056 (54) |
| **Age in years** – no. (%) |  |
| 70-74 yr | 7,894 (60) |
| 75-79 yr | 3,406 (26) |
| ≥80 yr | 1,831 (14) |
| **Race or ethnic group**<sup>a</sup> – no. (%) |  |
| White/Caucasian | 12,953 (99) |
| Other | 178 (1) |
| **Obese (BMI ≥ 30 kg/m<sup>2</sup>)**<sup>b</sup> (%) | 28 |
| **Current smoking** (%) | 4 |
| **Heart, stroke or vascular disease**<sup>c</sup> (%) | 0 |
| **Personal cancer history** (%) | 20 |
| Breast cancer history (%) | 4 |
| Colorectal cancer history (%) | 3 |

<sup>a</sup> Self-report.
<sup>b</sup> Obese was defined as body-mass index (weight in kilograms divided by the square of the height in meters) of ≥30.
<sup>c</sup> Baseline Characteristics of Participants in the ASPREE (ASpirin in Reducing Events in the Elderly) Study (22)

### PATHOGENIC VARIANTS IN MEDICALLY ACTIONABLE GENES

We detected 172 different pathogenic or likely pathogenic variants meeting ACMG/AMP guidelines (25) in 59 genes. Of these, 129 variants were in dominant genes, detected in 176 participants. We validated a representative sample of 10% of these 129 variants by Sanger sequencing (N=14), with 100% concordance. The highest number of pathogenic variants found in any one dominant gene was N=22 (*BRCA2* and *KCNQ1*)

We compared the prevalence of pathogenic variants in selected autosomal dominant genes in ASPREE versus the UK Biobank, a younger population of 49,960 adults aged average 58 years (26) (Table 2). We did this to determine if the prevalence of medically actionable variants was depleted in the healthy elderly, compared with a population-based cohort, irrespective of associated disease outcomes or age.

**Table 2.** Pathogenic Variants in Selected Autosomal Dominant Medically Actionable Genes in ASPREE versus the UK Biobank.

|  | UK Biobank<sup>a</sup><br>N=49,960 |  | ASPRE<br>N=13,131 |  |
| --- | --- | --- | --- | --- |
| **Average age – yrs (1<sup>st</sup>-3<sup>rd</sup> quartiles)** | 58 (45 to 71) |  | 75 (72 to 77) |  |
| **Female sex – %** | 55 |  | 54 |  |
| **Gene groups** | **No. of carriers** | **Carriers per 1000<sup>b</sup>** | **No. of carriers** | **Carriers per 1000<sup>b</sup>** |
| **ACMG59<sup>TM</sup> genes**<br>Autosomal dominant (N=55) | 994 | 20 | 176 | 13 |
| **Hereditary breast and ovarian cancer**<br>*BRCA1*<br>*BRCA2*<br>*BRCA1&BRCA2 Females* | 60<br>166<br>110 | 1<br>3.5<br>4 | 11<br>22<br>15 | 1<br>1.5<br>2 |
| **Lynch syndrome**<br>*PMS2*<br>*MSH6, MLH1, MSH2* | 59<br>78 | 1<br>1.5 | 12<br>8 | 1<br>0.5 |
| **Familial hypercholesterolemia (pathogenic)**<br>*LDLR, APOB, PCSK9* | 68 | 1.5 | 13 | 1 |
| **Romano-Ward long QT syndrome types 1,2, 3, Brugada**<br>*KCNQ1*<br>*KCNH2, SCN5A* | 55<br>61 | 1<br>1 | 22<br>17 | 1.5<br>1.5 |
| **Hypertrophic cardiomyopathy, Dilated cardiomyopathy**<br>*MYBPC3*<br>*MYH7, MYH11, LMNA, MYL3, TNNT2, TNNI3, GLA, MYL2* | 50<br>94 | 1<br>2 | 15<br>10 | 1<br>1 |
| **Arrhythmogenic right-ventricular cardiomyopathy**<br>*PKP2*<br>*DSC2, DSG2, DSP, TMEM43* | 55<br>98 | 1<br>2 | 12<br>10 | 1<br>1 |
| **Hereditary paraganglioma-pheochromocytoma syndrome**<br>*SDHB, SDHC, SDHD* | 33 | 0.5 | 5 | 0.5 |

<sup>a</sup>Van Hout et al, *Whole exome sequencing and characterization of coding variation in 49,960 individuals in the UK Biobank*, bioRxiv preprint first posted online Mar. 9, 2019
<sup>b</sup>Carrier rate per 1000 has been rounded to the nearest 0.5

Table 2 shows the number of variants detected in both studies across selected gene groups, and the prevalence of pathogenic variants per 1000 individuals (see Table S2 for individual gene frequencies). In ASPREE, pathogenic variants in medically actionable dominant genes were found in 1 in 75 healthy elderly adults (13 per 1000 participants). This was a lower carrier rate to 1 in 50 (20 per 1000) reported in the UK Biobank (26). Some genes had similar frequencies of pathogenic variants between the two studies. Notable differences were the lower prevalence of *BRCA2* variants observed in ASPREE, and lower prevalence of female *BRCA1/2* carriers (2 per 1000) versus the UK Biobank (4 per 1000).

We found no individuals carrying two or more variants in any autosomal recessive gene (homozygotes or compound heterozygotes). However, we found two individuals carrying pathogenic variants in two different dominant genes (*PMS2+KCNQ1* and *MYBPC3+LDLR* respectively).

### CLINICAL IMPACT OF PATHOGENIC VARIANTS OVER THE LIFETIME OF PARTICIPANTS

We compared the relative risk of associated disease in ASPREE variant carriers versus 12,936 non-carriers in the cohort, to mean age 79.5 years. In almost all cases, we found no statistically significant difference in relative risk of associated disease between pathogenic variant carriers and non-carriers, despite detection of pathogenic variants that infer high lifetime disease risk (Table S3). This was due to the high proportion of ASPREE participants who were free of the disease expected to be associated with the pathogenic variant at end of trial (Table S4).

The two exceptions were female *BRCA1/2* pathogenic variant carriers in ASPREE who were at increased risk of breast cancer versus non-carrier females (P=0.0003) and carriers of pathogenic variants in inherited cardiac disorder genes, who were at increased risk of rapid/sudden cardiac death versus non-carriers (P<0.0001). In the latter group, absolute risk was still low. Among 91 variant carriers, only two sudden cardiac deaths occurred in trial (97.8% associated disease-free). We compared the likelihood of death from any cause between pathogenic variant carriers and non-carriers to mean age 79.5 years, and found no statistically significant differences for any genes (Table S5).

### IDENTIFICATION OF RESILIENT INDIVIDUALS

To identify resilient individuals in the ASPREE population for medically actionable autosomal dominant genetic conditions, we excluded all pathogenic variant carriers in the cohort who had any symptoms of associated disease to mean age 79.5 years (for conditions where direct clinical phenotypes were available). This identified a total of 141 resilient candidates, who despite carrying pathogenic variants in dominant medically actionable genes, showed no apparent associated disease symptoms to the end of the trial (Table 3).

**Table 3.** Identification of Pathogenic Variant Carriers in ASPREE Free of Associated Autosomal Dominant Genetic Disease to Mean Age 79.5 Years (‘Resilience candidates’). We identified pathogenic variants in 55 autosomal dominant disease genes in 13,131 healthy elderly individuals enrolled in the ASPREE trial. We grouped related genes for autosomal dominant conditions and defined associated disease events in participant medical history (prior to study enrolment) and prospective disease diagnoses occurring during the ASPREE trial to median age 79.5 years.

|  |  | Pathogenic Variant Carriers in ASPREE |  |  |  |
| --- | --- | --- | --- | --- | --- |
| Mean age = 79.5 years |  | Total | With Associated Disease <sup>a</sup> | Without Associated Disease <sup>a</sup><br>'Resilient' | % Free of Associated Disease |
| **Hereditary breast and ovarian cancer syndrome (BRCA1, BRCA2), N=33 carriers** |  |  |  |  |  |
| **Cancers** | Breast (F) | 15 | 6 | 9 | 60% |
|  | Ovarian (F) | 15 | 0 | 15 | 100% |
|  | Prostate (M) | 18 | 2 | 16 | 89% |
|  | Melanoma | 33 | 3 | 30 | 91% |
|  | Pancreatic | 33 | 0 | 33 | 100% |
|  | **Total <sup>b</sup>** | **33** | **11** | **22** | **67%** |
| **Lynch syndrome (MLH1, MSH2, PMS2, MSH6) & Familial adenomatous polyposis (APC), N=21 carriers** |  |  |  |  |  |
| **Cancers** | Colorectal | 21 | 1 | 20 | 95% |
|  | Cervical/uterine (F) | 12 | 0 | 12 | 100% |
|  | Stomach | 21 | 0 | 21 | 100% |
|  | Gallbladder/bile duct | 21 | 0 | 21 | 100% |
|  | Ovarian (F) | 12 | 1 | 11 | 92% |
|  | **Total <sup>b</sup>** | **21** | **2** | **19** | **90%** |
| **Familial hypercholesterolemia (LDLR, APOB, PCSK9), N=13 carriers** |  |  |  |  |  |
| **CVD** | Myocardial infarction | 13 | 1 | 12 | 92% |
|  | Large artery atherosclerosis | 13 | 0 | 13 | 100% |
| **Deaths** | Coronary heart disease | 13 | 0 | 13 | 100% |
|  | **Total <sup>b</sup>** | **13** | **1** | **12** | **92%** |
| **Inherited cardiac disorders combined: Romano-Ward long QT syndrome (1,2,3), Brugada syndrome (KCNQ1, KCNH2, SCN5A), Hypertrophic, dilated cardiomyopathy (MYBPC3, LMNA, MYH11, MYH7, MYL3, TNNT2, GLA), Arrhythmogenic right-ventricular cardiomyopathy (PKP2, DSC2, DSG2, DSP, TMEM43), Catecholaminergic polymorphic ventricular tachycardia (RYR2), Marfan, Loeys-Dietz syndromes, and familial thoracic aortic aneurysms and dissections (FBN1), N=91 carriers** |  |  |  |  |  |
| **CVD** | Hospitalisation for heart failure | 91 | 1 | 90 | 99% |
| **Deaths** | Rapid/sudden cardiac | 91 | 2 | 89 | 98% |
|  | **Total <sup>b</sup>** | **91** | **3** | **88** | **97%** |
|  | **Grand Total <sup>b</sup>** | **158** | **17** | **141** | **89%** |

<sup>a</sup>To mean age 79.5 years, includes both associated disease events reported in personal medical history prior to study enrolment, and prospective events occurring during the ASPREE trial.
<sup>b</sup>Participants with >1 associated disease event were only counted once (F) = Female, (M) = Male

Of these 141 individuals, 22 carried pathogenic variants in genes associated with hereditary breast and ovarian cancer (*BRCA1/2*), 19 in genes associated with Lynch syndrome or familial adenomatous polyposis (*MLH1, MSH2, PMS2, MSH6, APC*), 12 in genes associated with familial hypercholesterolemia (*LDLR, APOB, PCSK9*), and 88 in genes associated with other inherited cardiac disorders or genetic arrythmias (*KCNQ1, KCNH2, SCN5A, MYBPC3, LMNA, MYH11, MYH7, MYL3, TNNT2, GLA, PKP2, DSC2, DSG2, DSP, TMEM43, RYR2, FBN1*).

## DISCUSSION

In the present study, we sequenced the DNA of 13,131 healthy elderly individuals and identified 141 apparent resilient carriers of pathogenic variants in genes which typically predispose to autosomal dominant conditions. Despite having a supposedly high genetic risk of disease, these individuals remained apparently unaffected until a mean age of 79.5 years. They were identified among a population of healthy elderly individuals meeting the strict eligibility criteria of the ASPREE trial (22). These elderly surviving pathogenic variant carriers are likely to be enriched for protective genetic factors.

There are important differences between resilience to childhood-onset and adult-onset genetic disorders. Many childhood-onset genetic disorders are autosomal recessive and fully penetrant, meaning when a copy of a pathogenic genetic variant is inherited from both parents, it almost always results in severe disease from an early age. Adult-onset conditions, by contrast, tend to be autosomal dominant, with only a single pathogenic variant required to infer risk or predisposition to disease. These conditions tend to occur later in life (25). Such conditions include hereditary breast and ovarian cancer (7), Lynch syndrome (9) and familial hypercholesterolemia (27). Most autosomal dominant conditions are not fully penetrant, meaning not all carriers of pathogenic variants develop the associated disease. Thus, a sub-set of pathogenic variant carriers for autosomal dominant conditions will remain unaffected, despite being at high genetic risk (28). These individuals (the lifelong unaffected) are less likely to be recruited in traditional genetic studies that ascertain participants on the basis of family or personal history of disease symptoms.

The pathogenic variants identified in this study might prompt medical interventions if detected at younger ages. For example, the risk management of pathogenic *BRCA1/2* variants might include risk-reducing bilateral mastectomy and bilateral salpingo-oophorectomy in younger women. Variants in other cancer-associated genes can also lead to risk-reducing surgeries or long-term high-risk cancer surveillance (29-31). Detection of pathogenic variants in inherited cardiac disorder genes might lead to implantation of automatic defibrillators (32). These interventions have substantial implications for both individuals and the healthcare system.

By comparison with the largest available population-based sequencing dataset (the UK Biobank) (26), we found the overall prevalence of pathogenic autosomal dominant variants to be higher (20 per 1000) than in ASPREE (13 per 1000). Differences in sequencing technology, variant curation or ethnic diversity may account for some variation in frequencies of pathogenic variants between the studies. However, the difference is also compatible with a survival disadvantage during the middle years of life.

In particular, the frequency of female *BRCA1/2* variant carriers in ASPREE was half that reported in the UK Biobank, reflecting a high penetrance of the *BRCA1/2* genes. Similarly, lower frequencies of variant carriers for the more penetrant Lynch syndrome genes (*MSH6, MLH1, MSH2*) was also observed, but not for the less penetrant (*PMS2*). For other genes, the carrier rate per 1000 was equivalent between ASPREE and the UK Biobank (e.g. *BRCA1, PMS2, MYBPC3, PKP2, SDHB, SDHC, SDHD*) and for occasional genes the carrier rate in ASPREE was higher (e.g. *KCNQ1, KCNH2, SCN5A*).

Our previous analysis of 589,306 genomes identified 13 rare exceptional ‘resilience candidates’, who despite having mutations that cause severe childhood-onset genetic disease, had no reported clinical symptoms to adulthood (13). Unfortunately, these individuals were identified through data aggregation from several different studies, meaning recontact or follow-up were not possible. Here, individuals have been identified who are resilient to adult-onset genetic disease by direct genomic screening of a healthy elderly cohort, accompanied by documented clinical outcome assessment, and with consent for re-contact (14).

The 141 resilience candidates identified now represent a unique sub-cohort for identification of protective genetic factors. Follow-up studies include deeper genomic characterization by whole genome sequencing and genome-wide genotyping, to determine the contribution of rare and common genetic factors towards risk modification (28). Contributions from environmental and lifestyle factors will also be examined through access to detailed longitudinal data collected by the ASPREE Longitudinal Study of Older Persons (33). Comparisons of ASPREE resilience candidates with other sample cohorts (e.g. case collections of those affected by early-onset disease for the same genes/conditions) are also being planned.

A strength of the current study is the detailed phenotyping with ascertainment and adjudication of outcome events during the 4.5 years of follow-up. The age of the cohort (all participants being above the average age of onset for most dominant conditions) has also facilitated the identification of those most likely to be resilient to disease development. Other strengths include the depth of sequencing coverage and stringency of variant curation applied, ensuring all pathogenic variant calls were of high confidence.

Our findings provide a scalable approach for more expansive genomic screening of the healthy elderly, to identify those resilient to adult-onset genetic disease. We identified resilience candidates in the healthy elderly at a rate of 1.07% (141 in 13,131 ASPREE participants). Projecting this to a larger sample population of 1 million healthy elderly individuals would estimate identification of over 10,000 resilience candidates. Such a population would provide the foundation for a statistical genetic study, with sufficient sample size to potentially identify protective genetic variants of genome-wide statistical significance (15). Such an approach is currently under consideration, involving both data mining of existing large population-based datasets for the healthy elderly, such as the UK Biobank (26), and direct screening of more community-based biobank cohorts of the healthy elderly, including the Statins in Reducing Events in the Elderly (STAREE), which is currently recruiting (14).

Our study also raises a clinically relevant question of whether there is an indicative age at which pathogenic variant carriers for dominant conditions outlive their genetic risk; that is, whether there is an age at which genetic risk and population risk for carriers of autosomal dominant pathogenic variants converge and become equal. Previous studies suggest the majority of clinical risk attributed to dominant pathogenic variants diminishes by age 70, especially for cancer (7, 9).

Limitations of our study include the ASPREE trial not originally being designed to analyse certain dominant genetic disease phenotypes. For example, electrocardiograms were not collected on all participants to detect cardiac abnormalities in pathogenic variant carriers for arrhythmia-related genes. We instead relied on hard study endpoints such as sudden cardiac death to define disease association in those genes. Technical limitations include a sequencing assay that was not capable of detecting pathogenic structural variants, including those in *MSH2* and other mismatch repair genes. This, in addition to the strict variant curation and filtering criteria, may have resulted in an under-estimation of pathogenic variant carriers identified. Some pathogenic variant carriers detected in ASPREE may have undergone risk-reducing surgery prior to study enrolment (including hysterectomy, for example, which removes ovarian cancer risk if ovaries are removed), and not reported this to ASPREE in the cancer screening history questionnaire (22). If this occurred in the absence of any cancer diagnosis, it could result in mis-attribution of resilience. Although unlikely, we cannot currently rule out this possibility without re-contacting the 41 resilient candidates for cancer-associated genes.

In summary, the study demonstrates that a proportion of healthy elderly carry pathogenic variants in medically actionable genes yet remain mostly free of associated disease. This provides a platform for the discovery of protective genetic factors in this uniquely resilient population, and a scalable method for identification of more resilient individuals in the future.

## Data Availability

n/a

## ACKNOWLEDGEMENTS

The ASPREE Healthy Ageing Biobank is supported by a Flagship cluster grant (including the Commonwealth Scientific and Industrial Research Organisation, Monash University, Menzies Research Institute, Australian National University, University of Melbourne); and grants (U01AG029824) from the National Institute on Aging and the National Cancer Institute at the National Institutes of Health, by grants (334047 and 1127060) from the National Health and Medical Research Council of Australia, and by Monash University and the Victorian Cancer Agency. We thank the trial staff in Australia and the United States, the participants who volunteered for this trial, and the general practitioners and staff of the medical clinics who cared for the participants.

## ETHICAL APPROVAL

This work was approved by the Alfred Hospital Human Research Ethics Committee (Project 390/15) in accordance with the National Statement on Ethical Conduct in Human Research (2007).

## AUTHOR CONTRIBUTIONS

PL and RS conceived the study; I.W, E.S and J.J.M directed the research; J.J.M, R.L.W, A.M.M, A.M.T, M.R.N and C.M.R directed the ASPREE trial; J.R, M.R, M.S, Y.C.W, R.C, B.A.T, D.D.B, P.A.J contributed to genetic analysis and variant curation; P.G and F.A.M contributed to adjudication of cancer outcomes; A.M.T was Chair of the cardiovascular outcomes adjudication committee; R.L.W, J.P, E.J.P, S.G.O, W.P.A, T.J.L, R.L.W, and J.J.M contributed to collection of biospecimens; R.S, R.C, D.S, T.A, M.S, Y.C.W and E.S contributed to panel design, DNA sequencing and variant calling; D.S, M.R and J.R contributed to the preparation of genetic data; J.E.L, S.G.O and R.L.W contributed to the preparation of phenotype data; D.D.B, F.A.M, P.A.J and I.W contributed to clinical interpretation of genetic variants; M.R and R.W contributed to statistical analyses; P.L wrote the manuscript, with editing from J.T, R.S, I.W and J.J.M. All authors reviewed the manuscript.

